# Grounding Language Models in Behavioral Science to Scale Physical Activity Interventions for Hispanic/Latinx Populations

**DOI:** 10.64898/2026.05.26.26354165

**Authors:** Sriya Mantena, Anders Johnson, Narayan Schuetz, Alexander Tolas, Samuel Montalvo, Juan Delgado-SanMartin, Mariana Ramirez-Posada, Lina Du, Sophia Zhang, Andy Duc Huynh, Marily Oppezzo, Abby C King, Paul Schmiedmayer, Allan Lawrie, Fatima Rodriguez, Euan A Ashley, Daniel Seung Kim

## Abstract

**Objective:** Hispanic/Latinx populations in the U.S. experience higher rates of chronic disease linked to physical inactivity, yet digital health interventions remain largely inaccessible to more than 16 million Hispanic/Latinx adults with limited English proficiency. While large language models (LLMs) offer scalable personalization, their use in non-English behavioral coaching is unexplored. This study introduces MHC-Coach-ES, a Spanish-language LLM fine-tuned on the Transtheoretical Model (TTM) of behavior change.

**Materials and Methods:** We fine-tuned Llama 3-70B-Instruct using a two-stage pipeline. First, the model was adapted to Spanish health and motivational language using a 2.21-million-token corpus. Second, it was instruction-tuned on 3,268 translated human written messages to align the model with the Transtheoretical Model (TTM) of Behavioral Change. We compared MHC-Coach-ES with Llama 3-70B-Instruct and translated human-expert messages using a forced-choice preference survey (N = 77) and blinded expert review (N = 2).

**Results:** Spanish-speaking participants significantly preferred MHC-Coach-ES messages over translated human-expert messages (81% preference, *P*<0.001). Linguistic analysis showed that MHC-Coach-ES produced more temporally anchored messages than the base model (65% vs. 20%), while maintaining readability. In blinded evaluation, clinical experts rated MHC-Coach-ES higher for alignment with Transtheoretical Model stages than human-expert messages (4.83 vs. 4.38 out of 5). The base model also outperformed translated expert messages across preference and expert ratings.

**Conclusions:** Generative AI can operationalize behavioral science frameworks in Spanish, offering a scalable approach to reducing health disparities. The strong performance of both MHC-Coach-ES and the base model highlights the promise of generative and personalized approaches over translation-based localization for theory-driven behavioral interventions.

## INTRODUCTION

Regular physical activity is a cornerstone in the prevention of many major chronic diseases, including cardiovascular disease, type 2 diabetes, dementia, and cancer^1,2^. Despite this, only a quarter of U.S. adults meet the physical guidelines recommended in the U.S. Department of Health & Human Services Physical Activity Guidelines for Americans^3^. Physical activity participation is significantly lower among Hispanic/Latinx populations than among non-Hispanic White adults, with only 23.5% of men and 18.0% of women meeting recommendations^4^. These patterns are compounded by structural and social determinants of health (e.g., household income, education), contributing to higher and worsening rates of metabolic syndrome in Spanish-speaking Hispanic/Latinx communities^5,6^. Consequently, there is an urgent need for culturally and linguistically tailored interventions that can sustain long-term increases in physical activity and help reduce differences in cardiovascular outcomes^7^.

Digital health interventions such as mobile apps, wearable smart devices, patient portals, and chatbots offer promise for delivering scalable, cost-effective physical activity promotion^8,9^. Yet, they consistently exclude individuals with limited English proficiency^10–13^. In 2024, approximately 25.7 million individuals in the United States had limited English proficiency, over 70% percent of whom was Hispanic or primarily Spanish-speaking^14^. Although 91% of Hispanic/Latinx adults own smartphones and frequently use them for health information^15^, many encounter persistent barriers, including limited translation support^16–18^. Despite their potential to support behavior change and improve long-term health outcomes, few commercial or research-backed health chatbots are available in Spanish^12,19–21^.

The linguistic gap extends to research and clinical trials, which form the scientific foundation of digital health technologies^22^. Recent analyses show that Hispanic/Latino participants are vastly underrepresented in digital health and behavioral intervention studies^23–25^. As a result, the evidence base guiding prevention strategies in cardiovascular and behavioral health may not generalize to the populations who could benefit most^7,13^. In studies that include Spanish-speaking participants, interventions are often limited to direct translations, failing to incorporate cultural norms, idiomatic expressions, or context-specific behavioral motivators^26^. Without in-depth engagement with linguistic and cultural variations, digital health efforts risk exacerbating existing disparities in care and outcomes^27,28^. In contrast, a small number of digital physical activity interventions targeting Hispanic/Latinx populations have used culturally and linguistically adapted eHealth tools and demonstrated significant increases in physical activity, comparable to outcomes achieved by trained human advisors^29,30^. These findings highlight the potential of culturally adapted digital interventions to effectively reach Hispanic/Latinx populations.

Beyond offering a Spanish-language interface, digital health tools must provide personalized support that adapts to users’ evolving needs. Generic, one-size-fits-all messages have repeatedly been shown to be ineffective for sustaining lasting behavioral change^31–34^. Until recently, delivering personalized coaching required rule-based logic or intensive human labor, constraining its scalability and impact^35^. Large Language Models (LLMs) offer a scalable alternative due to their ability to synthesize user data and generate context-specific messaging. Early proof-of-concept work that leverages off-the-shelf LLMs (e.g., OpenAI’s ChatGPT-4 and ChatGPT-3.5, Llama 2-7B-chat, and LaMDA) to generate motivational coaching messages shows that LLM responses are seen as more empathetic and actionable than human-written messages^36–39^. Yet these prototypes have two key limitations. First, they depend solely on ad-hoc prompt-engineering of general-purpose models that are not grounded in behavioral science theory, limiting the ability to systematically incorporate evidence-based mechanisms known to sustain long-term behavior change. Second, personalization is static, relying on generic motivational cues rather than dynamically adjusting to a user’s evolving stage of change, meaning they are neither equipped nor evaluated for longitudinal coaching.

A recent proof-of-concept study showed that fine-tuning a large language model on stage-labeled prompts enables generation of coaching messages aligned with the Transtheoretical Model (TTM) of Change^40^. The TTM defines five sequential stages of change, namely Precontemplation, Contemplation, Preparation, Action, and Maintenance, and has more than three decades of empirical support for guiding physical activity and other lifestyle interventions^41–43^. Although the prior work shows the feasibility of integrating LLMs with a rigorously validated behavioral framework, to our knowledge, all published behavioral science coaching systems have been developed and evaluated exclusively in English. This leaves nearly 26 million U.S. adults with limited English proficiency, most of whom prefer Spanish, without access to equally adaptive, theory-driven digital coaching.

Building on our prior English-language My Heart Counts LLM health coach (MHC-Coach)^44^, we developed MHC-Coach-ES, the first Spanish-language LLM system explicitly aligned with the Transtheoretical Model (TTM) for physical activity and exercise promotion. The model was trained using a two-stage pipeline that first adapts the model to Spanish-language health and motivational corpora, and then aligns its generation with TTM coaching messages derived from behavioral expert for each stage of behavioral change. We evaluate MHC-Coach-ES through a user preference survey, blinded expert assessment of effectiveness and stage alignment, and linguistic analysis of behaviorally relevant features. Together, these evaluations assess whether generative models can integrate language concordance, scalability, and behavioral-science alignment within a single framework for personalized physical activity and exercise support in multilingual populations.

## METHODS

This study consisted of five components. We first assembled two datasets: a Spanish Health & Motivation Corpus (SHMC) for domain adaptation and a translated Transtheoretical Model coaching dataset (TTM-T) for behavioral alignment (M.1). We then applied a two-stage fine-tuning pipeline to develop the MHC-Coach-ES model (M.2). Next, we compared linguistic and stylistic features across messages generated by multiple approaches (M.3). Finally, we evaluated model outputs using a user preference survey of Spanish-speaking participants (M.4) and blinded expert assessments of message effectiveness and behavioral-stage alignment (M.5).

### M.1 Training Corpus Assembly

#### M.1.1 Spanish Health & Motivation Corpus Construction

We curated a Spanish-language corpus for domain adaptation, hereafter referred to as the Spanish Health & Motivation Corpus (SHMC). SHMC was designed to reflect both native language fluency and domain-specific knowledge in health, motivation, and exercise. It includes research literature relevant to Spanish-speaking populations, educational materials on physical activity and exercise, and motivational public talks delivered in Spanish, to ground the model in culturally resonant language alongside evidence-based health guidance.

A total of 2.21 million tokens, measured using the Llama-3-70B SentencePiece tokenizer, were compiled from three document families (see **Supplemental Table 1**).

i. Peer-reviewed articles (n = 30) on exercise and physical activity promotion, cardiovascular risk reduction, and behavioral science coaching relevant to Spanish-speaking populations. English manuscripts were machine-translated into Spanish using the Google Translate v2 API, whereas native-Spanish texts were used as published.
ii. Consumer educational material, including government brochures, NGO pamphlets, and web-based coaching scripts in Spanish^105–116^. Additional evidence-based English brochures were translated to Spanish using the Google Translate v2 API to expand topical coverage^117–124^.
iii. **(iii)** Spanish TEDx transcripts were retrieved from the mTEDx^45^ corpus, a multilingual dataset of TEDx talk transcripts, and retained only if they contained at least two domain-specific keywords (e.g., salud, ejercicio, bienestar, dieta, hábitos saludables, prevención). These talks provide naturally motivational, culturally grounded Spanish that complements the more formal written sources. The full keyword list and citation ranges corresponding to each source category are provided in **Supplementary Table 2.**

After cleaning and sentence segmentation (Supplementary section S.1), the SHMC was used for the first stage of fine-tuning described in Section M.2.

#### M.1.2 Behavioral Science Dataset Construction

To align the model with the Transtheoretical Model (TTM) of behavior change, we used a coaching dataset originally written in English and subsequently translated into Spanish using the Google Translate v2 API. This dataset is hereafter referred to as the Translated TTM Coaching Dataset (TTM-T). This dataset comprised 3,268 motivational messages pairs written by 25 human peers and experts, including fitness coaches, behavioral psychologists, and health researchers^46^. Message writers were presented with scenarios of individuals in each stage of TTM behavioral change. Experts and peers were free to draw on any behavioral science strategy to motivate users in the five transtheoretical model stages: Precontemplation, Contemplation, Preparation, Action, and Maintenance. The resulting dataset contained approximately 215,000 tokens and served as supervised input for the second stage of fine-tuning described in Section M.2 (see **Supplemental Table 3**).

### M.2 Fine-Tuning

We adopted a two-stage fine-tuning strategy. In the first stage, the base Llama-3-70B-Instruct model was trained on the full 2.21 M-token SHMC to specialize it to domain-specific vocabulary and stylistic conventions. We selected Llama 3-70B^47^ due to its strong performance across reasoning and instruction-following benchmarks and its multilingual capabilities, including support for Spanish^48^.

Fine-tuning was performed using low-rank adaptation (LoRA), a parameter-efficient method for adapting large language models^49^. Hyperparameters were selected via grid search over learning rate, number of training epochs, LoRA rank, and batch size using a held-out validation set comprising 10% of the training data. The validation set was randomly sampled from SHMC at the document level. Model selection was based on minimizing validation loss. Full hyperparameter ranges and grid configurations are reported in **Supplementary Section S.1.2**. The final hyperparameter configuration consisted of four epochs with a batch size of 8 and LoRA adapters of rank 32 (α = 64) applied to all linear layers. Optimization followed a linear learning-rate schedule (initial learning rate 3 × 10⁻LJ with a warm-up period spanning the first 5% of training steps), complemented by gradient-norm clipping at 1.0 and no weight decay. Model performance was evaluated exclusively on downstream benchmarks disjoint from both the training and validation data.

In the second stage, we performed instruction-style fine-tuning on the TTM-T, comprising 3,268 stage-labeled coaching examples, to align generation with TTM structure. Hyperparameters were selected based on performance on a held-out validation set (**Section S.1.2**), yielding a final configuration of five epochs, the same LoRA topology used in Stage 1 (rank 32, α = 64, applied to all linear layers), a batch size of 8, and a peak learning rate of 2 × 10⁻□ with a warm-up stage over the first 3% of training steps. All other optimization settings followed those in Stage 1.

This two-stage fine-tuning strategy first adapted the model to Spanish-language health discourse using SHMC. The model was then instruction-tuned on TTM-T to align generation with the Transtheoretical Model (TTM). TTM-T consists of human-written coaching messages originally authored in English, machine-translated into Spanish, and labeled by TTM stage. This separation ensured that behavioral-stage alignment occurred atop a domain-relevant linguistic base (see **Figure 1**)^50,51^. The overall message generation and evaluation workflow, including generation methods in each evaluation, is summarized in **Figure 2**.

**Figure 1.**
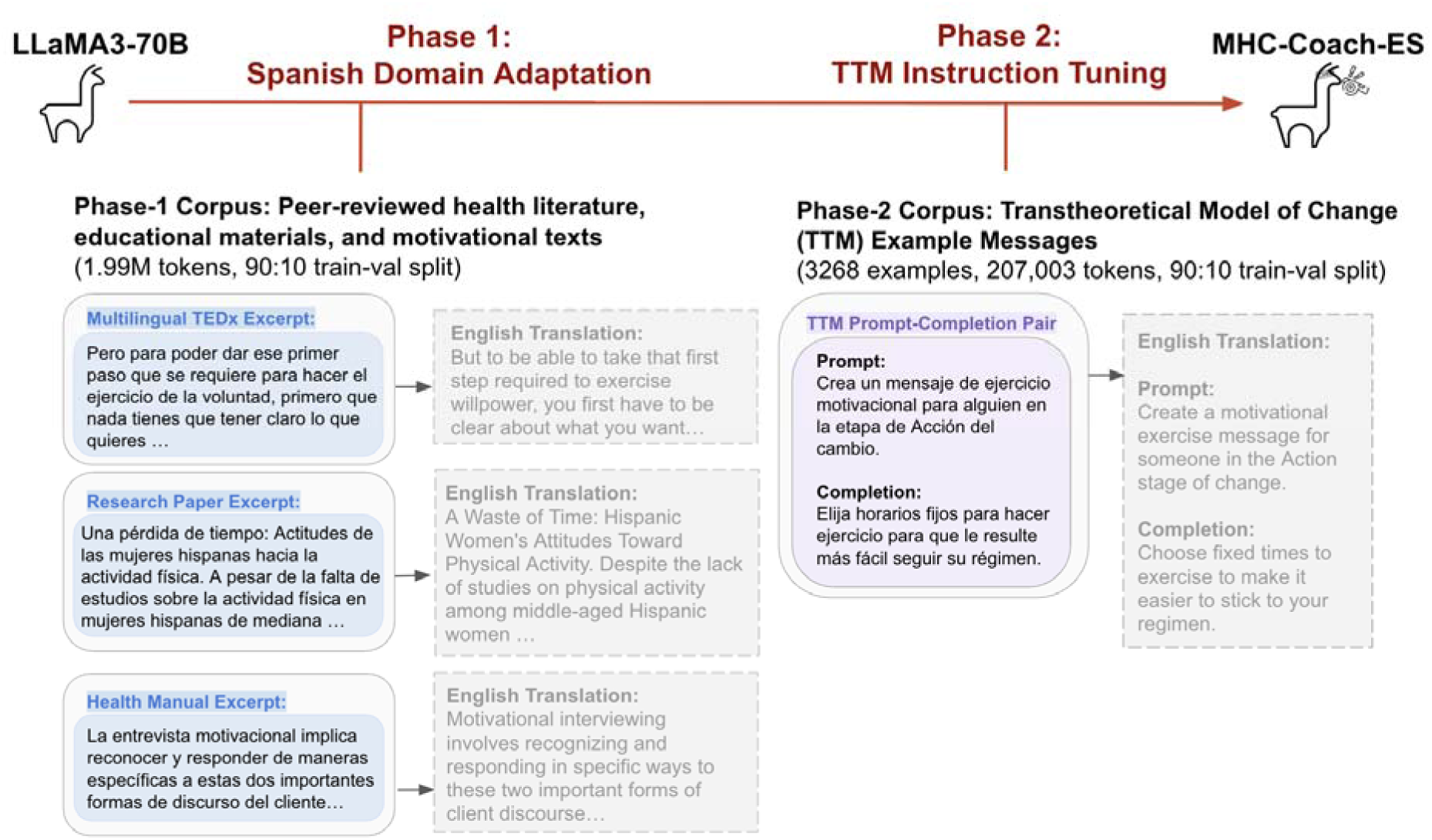
Overview of the MHC-Coach-ES training pipeline and evaluation. The Llama 3-70B-Instruct model was adapted in two stages: (1) domain adaptation using Spanish-language health and motivational corpora, and (2) instruction tuning on expert-authored coaching messages aligned with Transtheoretical Model (TTM) stages. The resulting model generates short motivational messages in Spanish, which are evaluated through user preference surveys and expert assessment.

**Figure 2.**
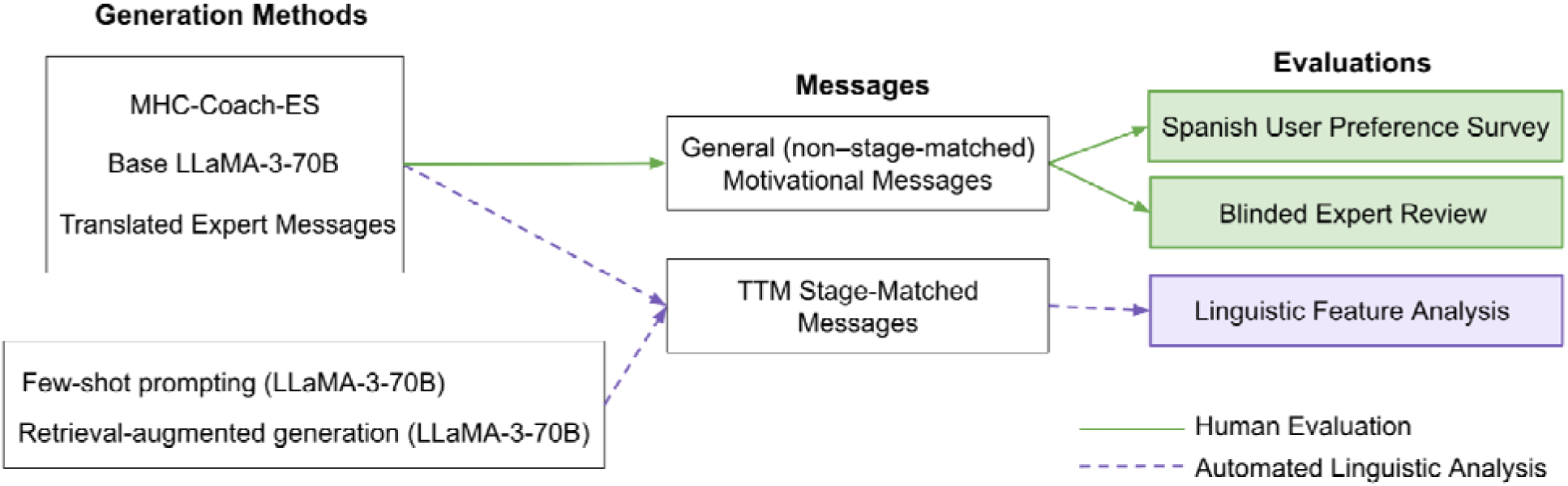
Message generation and evaluation pipeline. Five methods were used to generate Spanish motivational coaching messages. All generation techniques produced Transtheoretical Model (TTM) stage-matched messages, which were evaluated using automated linguistic feature analysis. MHC-Coach-ES, the base Llama-3-70B-instruct and translated human expert messages additionally generated general (non–stage-matched) motivational messages, which were included in the Spanish user preference survey and blinded expert review.

### M.3 Linguistic and Stylistic Features Across Generation Methods

#### M.3.1 Generation Methods

To compare MHC-Coach-ES with alternative generation approaches, we generated 100 stage-matched coaching messages using five methods: the base Llama-3-70B-Instruct model, few-shot prompting implemented with Llama-3-70B-Instruct using stage-specific example messages, retrieval-augmented generation (RAG)^52^ implemented on Llama-3-70B-Instruct using retrieved context from the TTM-T and SHMC corpora (embedded with the BAAI/bge-small-en-v1.5 encoder)^103,104^, the fine-tuned MHC-Coach-ES model, and expert-written messages sampled from TTM-T.

Across all model-based methods, decoding parameters were held constant, with a temperature of 0.8 and a top-p value of 0.7 to balance response diversity and coherence during generation. The same stage-specific base task prompt was used across methods, with method-specific context appended as appropriate. We generated four messages for each of the five TTM stages per method, yielding 20 messages per generation method. The prompt templates, retrieval procedures, and sampling specifications are provided in Supplementary Section S.2 (see **Supplemental Table 4**).

#### M.3.2 Message Feature Extraction

We conducted a linguistic comparison across methods using a set of predefined features that are associated with effective behavior change communication^53–57^, including word count, sentiment, lexical diversity, action-verb use, temporal anchoring, exclamation point usage, and readability. All messages were annotated using standard Spanish NLP tools (e.g., spaCy and TextBlob-ES). For each method, feature values were averaged across the 20 generated messages to obtain method-level estimates. Detailed feature definitions, preprocessing steps, and implementation details are reported in Supplementary Section S.3.

#### M.3.3 Statistical Comparison Across Methods

To test whether feature distributions differed by generation method, we analyzed seven linguistic features across messages (word count, sentiment, lexical diversity, action-verb use, temporal anchoring, exclamation point usage, and readability). Because feature distributions did not satisfy normality assumptions (Shapiro–Wilk test), we used the non-parametric Kruskal–Wallis test to assess the effect of the generation method on each feature. All statistical analyses were conducted in SciPy 1.13.

### M.4 User Preference Survey

#### M.4.1 Ethics

This study was reviewed and approved by Stanford University’s Research Compliance Office (IRB-75836). All procedures adhered to the principles of the Declaration of Helsinki and institutional ethical standards. Participants provided informed consent electronically through the Qualtrics platform and were informed that their responses would be anonymized and used solely for research purposes.

#### M.4.2 Survey Participants and Inclusion Criteria

Respondents were recruited through the Prolific^58^ online research platform in September 2025 and were eligible to participate if they were 18 years of age or older and self-identified as primary Spanish speakers. Eligible participants were directed to a Qualtrics survey, where they provided informed consent, completed a brief demographic questionnaire, and responded to the study questions. A total of 77 participants met all inclusion criteria and completed the full survey.

#### M.4.3 Survey

The survey consisted of an initial general motivation comparison block, followed by stage-specific comparison blocks assigned based on participants’ Transtheoretical Model (TTM) stage (**Figure 3**). In the general block, all participants were presented with the same pair of broadly motivational exercise messages: one generated by MHC-Coach-ES and one written by a human expert and translated into Spanish.

**Figure 3.**
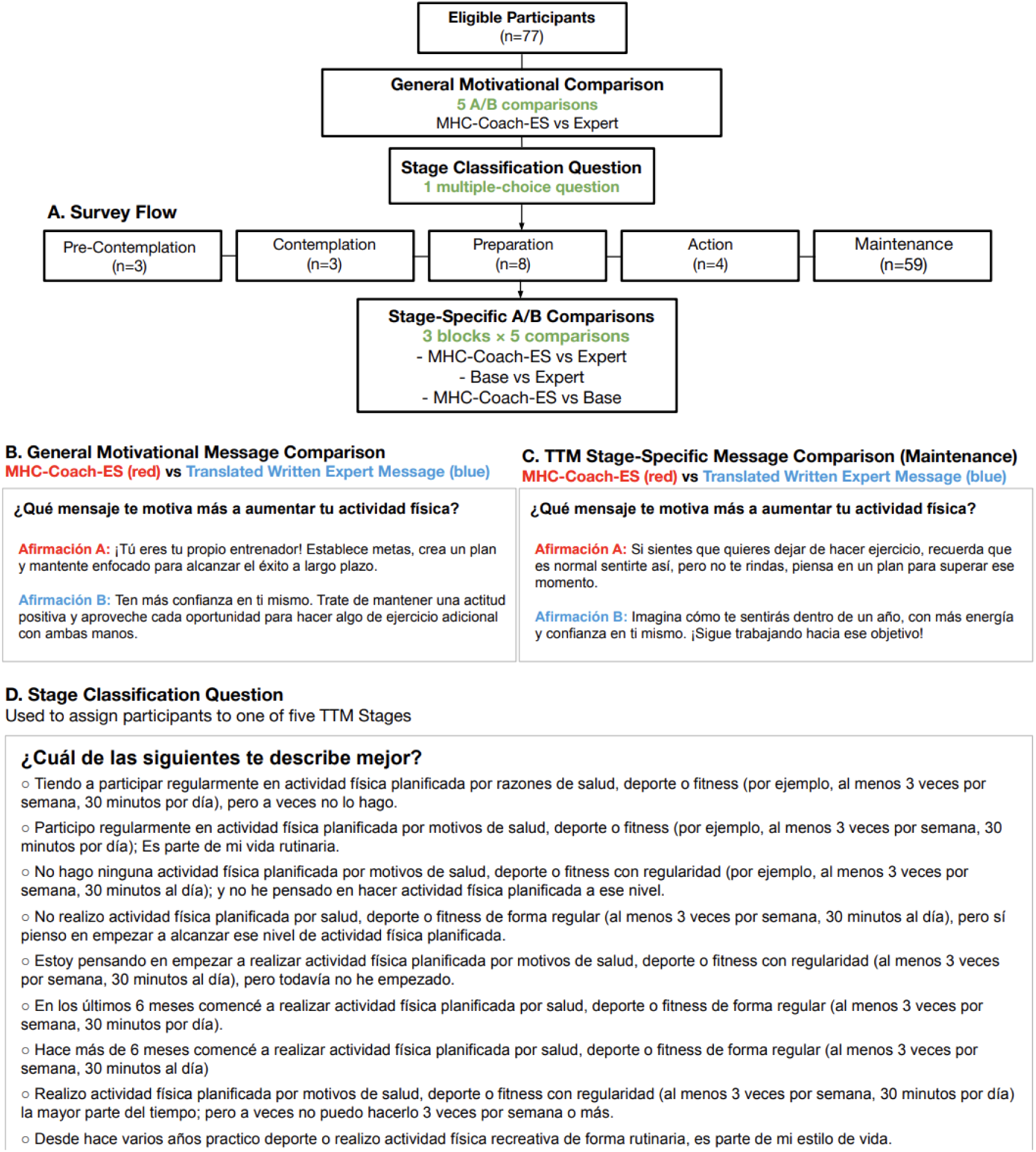
Survey design and message comparison procedure. (A) Overall survey flow. Spanish-speaking participants (n = 77) first completed a general motivational A/B comparison between MHC-Coach-ES and translated expert messages. Participants were then assigned to a Transtheoretical Model (TTM) stage of change using a multiple-choice item and completed stage-specific A/B comparisons corresponding to their assigned stage. These consisted of three comparison blocks (MHC-Coach-ES vs. expert, base Llama-3-70B-Instruct vs. expert, and MHC-Coach-ES vs. base), with five items per block. (B) Example general motivational message comparison. (C) Example stage-specific message comparison (Maintenance stage shown). (D) Stage-classification question used to assign participants to TTM stages. Panels B–D show representative examples; all participants completed the full set of comparisons described in panel A.

Participants’ TTM stage was determined using a brief multiple-choice item assessing exercise habits and mindset (**Figure 3**), mapping each individual to one of five stages: Precontemplation, Contemplation, Preparation, Action, or Maintenance. Based on this assignment, participants completed three stage-specific comparison blocks where they were presented messages tailored to their assigned stage of change: (1) MHC-Coach-ES vs. translated expert-written messages, (2) base Llama-3-70B-Instruct vs. translated expert-written messages, and (3) MHC-Coach-ES vs. the base model.

Each comparison block contained five forced-choice items. Participants were considered to prefer a message type if they selected it in at least three of five comparisons within a block, yielding a total of 20 forced-choice items per respondent (5 general and 15 stage-specific).

Human expert–written messages used across all comparisons were drawn from the same crowdsourced dataset used to fine-tune the model (the TTM-T), involving 25 human experts in fitness, behavior, and health psychology^59^. These messages, drawn from TTM-T, were originally authored in English and translated to Spanish using the Google Cloud Translation API v2. To ensure comparability with model-generated outputs, which were prompted to produce brief coaching messages of approximately 20 words, we excluded very short expert responses and randomly sampled messages from those longer than the dataset’s median length. All participants were shown the same set of message pairs, enabling direct evaluation of whether model outputs matched expert-quality messages from the training distribution.

### M.5 Expert Evaluation of Messages

Message annotations and ratings were performed by two domain experts: one Spanish-speaking physician with clinical experience in behavior-change counseling, and one Spanish-speaking behavioral science specialist. Before rating, both experts received standardized information on the TTM, including concise stage definitions and an evaluation rubric, to ensure consistent application of rating criteria. Each expert independently evaluated MHC-Coach-ES–generated messages and human expert-written messages sampled from the TTM-T dataset, in a blinded fashion (see Methods Section M.4.3 for message selection). Each message was rated on a 5-point Likert scale along two criteria: (1) the anticipated effectiveness for increasing physical activity and exercise, and (2) the alignment to a specific TTM stage of change. Inter-rater reliability was assessed using the intraclass correlation coefficient (ICC).

## RESULTS

### R.1 Linguistic Feature Analysis

To compared the outputs of MHC-Coach-ES with other generation strategies and expert-written messages, we analyzed seven linguistic and stylistic features that are commonly associated with effective behavior change communication: word count, sentiment, action-verb frequency, temporal reference, lexical diversity (type–token ratio), exclamation point usage, and readability^53–57^. For each feature, values were averaged across the 20 messages produced by each method (four messages per TTM stage).

Descriptive analyses revealed systematic stylistic differences across generation methods (**Table 1**). MHC-Coach-ES produced the highest rate of temporally anchored messages (e.g., “en los próximos 30 días,” “después de seis meses”), with 65% of generated messages containing explicit temporal references, and exhibited the highest average readability score, while maintaining a message length comparable to other model-based approaches. Action-verb use in MHC-Coach-ES (mean = 1.4 per message) was similar to the base model and exceeded that of expert-translated messages.

**Table 1.** Linguistic and stylistic feature comparisons across generation methods. Mean values are reported for each feature across five generation methods (N = 20 messages per method). Features include word count (WC), sentiment index (Sent), action-verb count (AV; mean number of action verbs per message), temporal referencing (Temp; proportion of messages containing explicit temporal cues), type–token ratio (TTR; lexical diversity), exclamation point usage (Exc), and Fernández–Huerta readability (FH; higher values indicate easier readability). The final two columns summarize inferential results from Kruskal–Wallis tests followed by Dunn post-hoc comparisons with Holm correction. Strongest significant pair denotes the pair of methods exhibiting the largest statistically significant difference for a given feature; Mean difference reports the magnitude of this difference. A dash indicates that no statistically significant between-method differences were detected for that feature. Detailed feature definitions are provided in Supplementary Section S.3.

| Feature | Base Model | MHC-Coach-ES | Few-shot | RAG | Expert | Strongest Significant Pair | Mean Difference |
| --- | --- | --- | --- | --- | --- | --- | --- |
| Word Count (WC) | 22.2 | 22.9 | 20.95 | 20.3 | 14.1 | Base Model > Expert | 8.1 |
| Sentiment (Sent) | 0.63 | 0.68 | 0.76 | 0.68 | 0.46 | - | - |
| Action Verb Count (AV) | 1.45 | 1.4 | 1.1 | 1.8 | 0.65 | RAG > Expert | 1.15 |
| Temporal Reference (Temp) | 0.2 | 0.65 | 0.55 | 0.45 | 0.35 | MHC-Coach-ES > Base Model | 0.45 |
| Type–Token Ratio (TTR) | 0.92 | 0.91 | 0.91 | 0.95 | 0.95 | - | - |
| Exclamation Point Count (Exc) | 1.7 | 1.5 | 0.7 | 0.8 | 0.4 | Base Model > Expert | 1.3 |
| Readability (FH) | 90.6 | 94.14 | 89.55 | 86.94 | 90.82 | - | - |

Retrieval-augmented generation (RAG), implemented on the base Llama-3-70B-Instruct model with retrieved context from TTM-T and SHMC, produced the highest action-verb frequency (mean = 1.8 per message) and high lexical diversity, but showed low temporal anchoring (45% of messages) and the lowest readability scores. The base Llama-3-70B-Instruct model, using only the shared base prompt, demonstrated the weakest temporal anchoring (20% of messages) and the highest exclamation point usage, alongside moderate sentiment. Few-shot prompting, implemented on the same base model with stage-specific message examples appended to the prompt, yielded the highest sentiment scores and intermediate temporal anchoring, but lower action-verb frequency. Lexical diversity was uniformly high across all generation methods.

To assess whether these stylistic differences were statistically significant, we conducted Kruskal–Wallis tests across generation methods for each linguistic feature. Significant method effects were observed for word count (H = 38.48, p < 0.001), action-verb frequency (H = 11.28, p = 0.024), temporal referencing (H = 9.80, p = 0.044), and exclamation point usage (H = 15.03, p = 0.005). No significant differences were detected for sentiment (H = 2.02, p = 0.732), lexical diversity (H = 6.61, p = 0.158), or readability (H = 3.48, p = 0.48). Post hoc comparisons indicated that differences in word count and exclamation point usage were primarily driven by the base model producing longer and more emphatic messages than expert-translated texts. Higher action-verb frequency reflected retrieval-augmented generation producing more behavioral directives than the expert baseline. Differences in temporal referencing were most pronounced between the base and fine-tuned models, with MHC-Coach-ES exhibiting substantially greater use of explicit time cues. Full statistical results are reported in **Table 1**.

### R.2 User Preference Survey

Of the 90 individuals who began the survey, 77 met the eligibility criteria (≥18 years, Spanish-speaking) and provided complete responses, forming the final analytic sample.

The sample was predominantly female (64.9%), with 32.5% identifying as male and 2.6% as non-binary or a third gender. Participants were largely in young to mid-adulthood: one-third were aged 20–29 (33.8%) and another third 30–39 (35.1%), with smaller proportions aged 40–49 (14.3%), 50–59 (10.4%), and 60 or older (6.5%). Most respondents identified as Hispanic, Latino, or of Spanish origin (79.2%), followed by multiethnic (9.1%), White (6.5%), Black/African American (2.6%), Asian (1.3%), and prefer-not-to-say (1.3%). Nearly half of participants reported Spanish as their first language (49.4%), while others reported English (28.6%) or Spanish–English bilingualism (22.1%); all participants were proficient Spanish speakers. Full demographic characteristics are provided in **Table 2**.

**Table 2.** Participant Demographics: Summary of the study population’s demographic characteristics, including age, gender, ethnicity (with the option to select multiple ethnicities), and native language.

| Category | Subcategory | Count | % |
| --- | --- | --- | --- |
| Gender | Male | 25 | 32.5% |
|  | Female | 50 | 64.9% |
|  | Non-Binary / Third Gender | 2 | 2.6% |
|  | Prefer not to say | 0 | 0.0% |
| Age Group | 10–19 | 0 | 0.0% |
|  | 20–29 | 26 | 33.8% |
|  | 30–39 | 27 | 35.1% |
|  | 40–49 | 11 | 14.3% |
|  | 50–59 | 8 | 10.4% |
|  | 60+ | 5 | 6.5% |
| Ethnicity | Hispanic, Latino, or Spanish origin | 61 | 79.2% |
|  | Multiethnic | 7 | 9.1% |
|  | Black or African American | 2 | 2.6% |
|  | White | 5 | 6.5% |
|  | Asian | 1 | 1.3% |
|  | Prefer not to say | 1 | 1.3% |
| Native Language | Spanish | 38 | 49.4% |
|  | English | 22 | 28.6% |
|  | Spanish and English | 17 | 22.1% |

When shown general motivational messages that were not tailored to a stage of change, participants preferred MHC-Coach-ES over expert-translated messages (78% selection of MHC-Coach-ES; p = 1.6×10⁻□), indicating a overall preference for the MHC-Coach-ES messages in the general context (**Table 3**).

**Table 3.** Percentage of respondents selecting the left-hand option in each A/B comparison. Values are reported as counts and percentages of participants preferring each message type within each comparison. P-values are derived from χ² goodness-of-fit tests (1 degree of freedom) against a 50% null preference and are reported only for aggregated and sufficiently powered comparisons. The general comparison includes all participants, whereas stage-specific results are stratified by Transtheoretical Model (TTM) stage. Dashes indicate comparisons for which statistical testing was not performed due to limited sample size.

| Stage (n) | MHC-Coach-ES<br>> Expert N (%) | Chi-Square<br>P-Value | Base > Expert<br>N (%) | Chi-Square<br>P-Value | MHC-Coach-ES<br>> Base N (%) | Chi-Square<br>P-Value |
| --- | --- | --- | --- | --- | --- | --- |
| General Motivational<br>Messages |  |  |  |  |  |  |
| All ( n= 77) | 60 (78%) | 1.60E-05 | — | — | — | — |
| Stage-Specific<br>Messages |  |  |  |  |  |  |
| Precontemplation (n = 3) | 3 (100%) | — | 2 (67%) | — | 2 (67%) | — |
| Contemplation (n = 3) | 3 (100%) | — | 3 (100%) | — | 1 (33%) | — |
| Preparation (n = 8) | 6 (75%) | — | 7 (88%) | — | 5 (63%) | — |
| Action (n = 4) | 3 (75%) | — | 3 (75%) | — | 1 (25%) | — |
| Maintenance (n = 59) | 47 (80%) | 5.20E-06 | 52 (88%) | 4.67E-09 | 35 (59%) | 0.15 |
| Total (n = 77) | 62 (81%) | 1.60E-05 | 67 (87%) | 8.30E-11 | 44 (57%) | 0.21 |

We examined A/B choices within stage-specific pairings (**Table 3**). Preferences were not evenly distributed across stages, with the Maintenance subgroup comprising most respondents (n=59), while other stages were sparsely represented (e.g., Precontemplation n=3, Contemplation n=3, Action n=4).

When aggregated across stages, respondents showed a strong preference for model-generated messages relative to expert-translated messages and a more modest preference for MHC-Coach-ES over the base model. In the two stages with the most respondents, Maintenance (n = 59) and Preparation (n = 8), 80% and 75% of participants, respectively, preferred MHC-Coach-ES over expert translations, while 88% in both stages preferred the base model over expert translations. We observed similar preference patterns in earlier stages, though the limited data restricts these results to a descriptive analysis. Descriptive analyses did not reveal consistent differences in preferences across demographic subgroups (e.g., gender, age, or education level).

Aggregated across stages, 81% of participants preferred MHC-Coach-ES to expert-translated messages (p < 0.001), whereas 87% preferred the base model to expert translations (p < 0.001). Preference for MHC-Coach-ES over the base model was more modest (57%) and did not reach statistical significance (p = 0.21). Full per-stage and aggregate results are reported in **Table 3**.

### R.3 Expert Message Rating

One physician and one behavioral science expert evaluated messages from the base model, MHC-Coach-ES, and the human expert corpus on two dimensions: perceived effectiveness for increasing physical activity/excercise and alignment with the appropriate TTM stage of change. Ratings were averaged across two independent expert evaluators who assessed each message in a blinded fashion. Inter-rater reliability was low for both perceived effectiveness (intraclass correlation coefficient [ICC] = 0.18) and stage alignment (ICC = −0.03), indicating variability in how evaluators applied the rating criteria. Accordingly, expert ratings are presented as descriptive averages across the two raters.

Across both dimensions, the two LLM-based systems received higher expert ratings on average than the expert-translated messages. For perceived effectiveness, both the base model and MHC-Coach-ES received similarly high mean ratings on a 5-point likert scale (mean rating = 4.68), exceeding those assigned to expert-translated messages (mean rating = 3.90). For stage alignment, expert ratings again favored the LLM-based systems, with MHC-Coach-ES receiving the highest mean score (mean rating = 4.83), closely followed by the base model (mean rating = 4.80), compared with expert-translated messages (mean rating = 4.38).

Differences between the base model and MHC-Coach-ES were most apparent in earlier stages of change. In the Precontemplation stage, experts rated MHC-Coach-ES as more effective than the base model (mean rating = 4.00 vs. 3.75). In the Contemplation stage, the two systems received equivalent effectiveness ratings (mean rating = 4.63 for both), although MHC-Coach-ES demonstrated stronger alignment with the assigned TTM stage (mean rating = 5.00 vs. 4.38). Detailed per-stage expert ratings are reported in **Table 4**.

**Table 4.** Expert ratings of message effectiveness and stage alignment across Transtheoretical Model (TTM) stages. Ratings were provided on a 5-point Likert scale (1 = very low, 5 = very high). Both the base and MHC-Coach-ES models consistently outperformed expert-authored messages on perceived effectiveness and stage match, with MHC-Coach-ES displaying a slightly higher stage match.

| Stage | MHC-Coach-ES Effectiveness | Base Model Effectiveness | Expert Effectiveness | MHC-Coach-ES Match | Base Model Match | Expert Match |
| --- | --- | --- | --- | --- | --- | --- |
| Precontemplation | 4.00 | 3.75 | 3.63 | 4.75 | 4.75 | 4.00- |
| Contemplation | 4.63 | 4.63 | 4.25 | 4.38 | 5.00 | 4.25 |
| Preparation | 4.75 | 5.00 | 3.75 | 5.00 | 4.5 | 4.25 |
| Action | 5.00 | 5.00 | 3.88 | 5.00 | 5.00 | 4.38 |
| Maintenance | 5.00 | 5.00 | 4.00 | 5.00 | 4.75 | 5.00 |
| **Overall** | **4.68** | **4.68** | **3.90** | **4.83** | **4.80** | **4.38** |

## DISCUSSION

Digital health coaching offers a powerful lever for the personalized prevention of chronic disease at scale^60^. This study extends the application of generative AI–driven messaging to Spanish-speaking users, a population disproportionately affected by cardiovascular disease structural barriers to prevention, including reduced access to culturally and linguistically appropriate health resources^61^. To help address these disparities, we developed and evaluated the first LLM-based physical activity coach fine-tuned on the TTM of behavior change, specifically adapted for Spanish-speaking populations. Earlier research has demonstrated that culturally adapted, Spanish-language digital interventions can increase physical activity and exercise in diverse groups^62–64^, but often these programs have relied on manually scripted, expert-authored content, limiting their reach^60^. Our approach introduces a generative system capable of producing idiomatic, stage-specific coaching messages tailored to behavioral readiness at scale.

This study evaluates whether large language models can be adapted to deliver theory-driven behavioral coaching in Spanish. Using a two-stage, fine-tuning pipeline, we aligned a model with Spanish-language health discourse and the TTM, a framework with strong empirical support for behavior change (MHC-Coach-ES)^41,43,65,66^. We compared this approach to alternative generation techniques, including the base Llama-3-70B instruct model, few-shot prompting, and retrieval-augmented generation (RAG), to generate behaviorally aligned messages. MHC-Coach-ES demonstrated significant advantages in linguistic features such as temporal anchoring, action-verb usage, and readability. To our knowledge, this study represents the first two-stage fine-tuning pipeline for behavioral science-grounded multilingual health coaching.

Interestingly, our findings show that out-of-the-box LLMs already perform well in generating behaviorally aligned health coaching messages. Both linguistic analyses and expert clinician ratings showed that the base model achieved high effectiveness and stage alignment scores, surpassing expert-written messages and often comparable to the fine-tuned system. This suggests that large foundation models already possess strong implicit behavioral reasoning capabilities. Fine-tuning, therefore, serves to constrain generation according to validated behavioral theory and to adapt model behavior to specific health-coaching contexts and populations.

Direct evaluation by Spanish-speaking participants showed that, although MHC-Coach-ES was often favored, its advantage over the base model was less pronounced than suggested by linguistic feature analysis. This divergence raises a key question about how we define and measure effective coaching. While frameworks like the TTM provide a useful structure for modeling behavior change, our findings suggest that preference is influenced by features that extend beyond readability, timeliness, and actionability. Smaller studies in demographically specific groups have highlighted the role of social support, family obligations, and culturally salient norms in Spanish-speaking populations compared with the contexts where these frameworks were developed^67,68^; however, these insights remain fragmented and not standardized in evaluation practices. Importantly, AI provides a unique opportunity to both evaluate and discover motivational features by tailoring interventions to specified demographics and analyzing feedback at scale. This approach could support the creation of Spanish benchmarks for AI models and, more broadly, expand behavioral science frameworks to become more fine-grained and responsive across diverse populations.

Embedding MHC-Coach-ES into the future My Heart Counts smartphone application provides a pathway to operationalize this alignment^69^. By collecting ongoing data on user responses and preferences, the system can refine evaluation criteria to better match effectiveness on Spanish-speaking populations. Additionally, emerging AI methods, including reinforcement learning from human feedback^70^, memory-augmented architectures^71^, and multimodal architectures^72^, can further integrate real-time activity data, health history, and regional linguistic preferences to support deeper personalization of coaching.

The two-stage fine-tuning pipeline we developed is model-agnostic and compatible with open-source LLMs of varying sizes. This enables flexible deployment, including on-device implementations that minimize latency and preserve user privacy by keeping their data on local devices. Additionally, unlike prompting-based methods, fine-tuning eliminates the need for runtime context engineering, simplifying integration and lowering latency.

Several limitations should be noted. First, participants in the Maintenance stage were over-represented and nearly 70% of participants were under the age of 40. This skew toward individuals in later stages of change, who are already motivated and behaviorally engaged, as well as toward younger adults, may constrain the generalizability of findings across the full TTM spectrum and to older populations. Second, some educational materials and expert comparator messages were machine-translated to Spanish for training dataset construction and not systematically reviewed by native speakers. Model outputs were instead validated by Spanish-speaking participants and clinicians during evaluation, enabling output-level empirical assessment within a scalable framework. This design enabled empirical validation of model outputs in the target population while supporting a scalable experimental pipeline based on translated expert data. Moreover, AI-generated messages were not tailored to cultural or regional backgrounds (e.g., Mexican, Colombian, Peruvian), which may influence how coaching messages are received and interpreted. While our findings offer promising signals of motivational alignment, future work is needed to evaluate downstream behavioral and cardiovascular impacts. Finally, while MHC-Coach-ES was fine-tuned on scientific health data to enhance accuracy, we did not conduct a systematic evaluation of hallucinations. Future work should address hallucination risks in multilingual settings, where cross-language generalization may introduce language-specific errors, particularly for less-resourced dialects^73^.

## CONCLUSION

This study demonstrates how behavioral-science grounded, language-specific fine-tuning can move digital health coaching beyond scripted interventions toward a new generation of adaptive, culturally responsive systems. We show that two-stage fine-tuning strengthens motivational features beyond what base models, prompting approaches, or translated expert messages can deliver, and is preferred by Spanish speakers as compared to human expert prompts. These findings establish not only the first Spanish fine-tuning pipeline for behavior change but also a general framework for adapting LLMs to multilingual populations. In the future, integrating MHC-Coach-ES into the My Heart Counts smartphone application will allow evaluation of its impact on engagement, adherence, and cardiovascular outcomes. By coupling scalable generative technology with behavioral science, this work points toward digital health coaching that is both theoretically aligned and culturally attuned.

## Data Availability

Our training dataset was constructed from four sources. First, we compiled 30 peer-reviewed articles. A subset of these were open-access publications, while the remainder were accessed through an academic subscription. Full texts were used exclusively for academic research under fair-use provisions and are not redistributed or publicly released. Second, we used Spanish and Spanish-translated physical-activity educational materials, all of which are open source; the aggregated transcripts used for model training are available on GitHub (https://github.com/SriyaM/MHC-Coach/tree/main). Third, we incorporated Spanish transcript excerpts from the mTEDx corpus, which are also publicly available on GitHub. Finally, we included messages written by peers and experts based on the Transtheoretical Model from a separate 2017 study. These messages, authored by human experts and crowd contributors, were translated into Spanish for this work and are not publicly available. Researchers interested in accessing this dataset may contact the author of the original study.
The preference data collected and analyzed in this study are available on GitHub and have been fully anonymized to protect participant privacy. The full set of messages generated by all five techniques (MHC-Coach-ES, base Llama-3-70B-instruct, few-shot prompting, RAG, and expert) and used for the linguistic and qualitative analyses is also included in the GitHub repository.

## ACKNOWLEDGEMENTS

D.S.K. discloses support for the research of this work from the Wu Tsai Human Performance Alliance (Clinician-Scientist Fellow), the Stanford Center for Digital Health (Digital Health Scholar), the Pilot Grant from the Stanford Center for Digital Health, the Robert A. Winn Excellence in Clinical Trials Career Development Award, the National Institutes of Health (NIH 1L30HL170306), the American Heart Association Career Development Award (25CDA1436622), and the American Diabetes Association Pathway to Stop Diabetes Initiator Award (7-25-INI-11).

F.R. discloses support for the research of this work from the NIH National Heart, Lung, and Blood Institute (R01HL168188; R01HL167974; R01HL169345), the American Heart Association/Harold Amos Medical Faculty Development Program, and the Doris Duke Foundation (Grant #2022051).

A.C.K. discloses support for the research of this work from the National Institutes of Health (5R01AG07149002; 1R44AG071211) and a seed grant from the Stanford Institute for Human-Centered Artificial Intelligence.

A.L. discloses support for the research of this work from the Imperial British Heart Foundation Research Excellence Award (RE/24/130023) and the British Heart Foundation Programme Grant (RG/F/25/110167).

## CODE/DATA AVAILABILITY

Our training dataset was constructed from four sources. First, we compiled 30 peer-reviewed articles^2,42,74–101^. A subset of these were open-access publications, while the remainder were accessed through an academic subscription. Full texts were used exclusively for academic research under fair-use provisions and are not redistributed or publicly released. Second, we used Spanish and Spanish-translated physical-activity educational materials, all of which are open source^105–124^; the aggregated transcripts used for model training are available on GitHub (https://github.com/SriyaM/MHC-Coach/tree/main). Third, we incorporated Spanish transcript excerpts from the mTEDx corpus^45^, which are also publicly available on GitHub. Finally, we included messages written by peers and experts based on the Transtheoretical Model from a separate 2017 study^102^. These messages, authored by human experts and crowd contributors, were translated into Spanish for this work and are not publicly available. Researchers interested in accessing this dataset may contact the author of the original study.

The preference data collected and analyzed in this study are available on GitHub and have been fully anonymized to protect participant privacy. The full set of messages generated by all five techniques (MHC-Coach-ES, base Llama-3-70B-instruct, few-shot prompting, RAG, and expert) and used for the linguistic and qualitative analyses is also included in the GitHub repository.

## AUTHOR CONTRIBUTIONS

Daniel Seung Kim and Sriya Mantena conceived the core ideas of the study. Sriya Mantena developed the computational methodology. Sriya Mantena, Anders Johnson, Alexander Tolas, Lina Du, Sophia Zhang, and Andy Duc Huynh designed the survey. The theoretical framework of this work was informed by Narayan Schuetz, Marily Oppezzo, Abby C King, Allan Lawrie, Fatima Rodriguez, and Paul Schmiedmayer. Samuel Montalvo, Juan Delgado-SanMartin, Fatima Rodriguez, and Mariana Raimrez-Posada provided linguistic expertise on the Spanish translations. All authors contributed to the technical discussions and writing of the manuscript. Daniel Seung Kim and Euan A Ashley supervised the research.

