## Supplemental Materials for "Grounding Language Models in Behavioral Science to Scale Physical Activity Interventions for Hispanic/Latinx Populations"

**SUPPLEMENTARY INFORMATION**

**Supplementary Table 1.** Composition of the Spanish Health & Motivation Corpus (SHMC) used for Phase 1 training of MHC-Coach-ES. The corpus totaled 2.21 million tokens after preprocessing, measured using the LLaMA-3-70B SentencePiece tokenizer. Sources include peer-reviewed research articles, informational brochures and pamphlets, and Spanish-language TEDx transcripts. Articles include both native Spanish manuscripts and machine-translated English texts; brochures include translated English health guides. TEDx transcripts were retained only if they contained at least two health-related keywords (e.g., salud, ejercicio; full keyword list provided in Supplementary Table 1). The corpus was partitioned into 90% training and 10% validation splits.

| **Source category** | **Train tokens (BPE)** | **Validation tokens (BPE)** | **Total tokens (BPE)** | **Share of corpus**  **(%)** |
| --- | --- | --- | --- | --- |
| Peer-reviewed articles | 451,719 | 50,869 | 502,588 | 22.77% |
| Informational brochures & pamphlets | 757,129 | 80,974 | 838,103 | 37.97% |
| TEDx transcripts | 780,343 | 86,106 | 866,449 | 39.26% |
| Corpus total | 1,989,191 | 217,949 | 2,207,140 | 100.00% |

**Supplementary Table 2:** Source categories and citation ranges for documents included in the Spanish Health & Motivation Corpus (SHMC). This table lists the source categories used to construct the SHMC and provides the corresponding citation ranges for each category. Sources include peer-reviewed research articles, consumer-facing educational materials, and Spanish-language TEDx transcripts. TEDx transcripts were retained only if they contained at least two of the listed keywords. English-language sources were machine-translated into Spanish using the Google Translate v2 API, while native Spanish texts were used as published.

| **Source category** | **Content and References** | **Keyword filters** |
| --- | --- | --- |
| Peer-reviewed articles | 30 peer-reviewed research articles on physical activity, cardiovascular health, and behavioral science coaching[^2,42,74–101^](https://paperpile.com/c/jexwV6/5Ovi+A871+PISL+W1rt+nmvC+uDWn+ZC5E+4vV3+aGNH+OhHg+tdIT+9Vu8+8gFP+9H69+iWY3+KHC2+CNYY+9hEw+zXl1+T4tL+HjHhi+yQLye+r3QW+Rt45+hiLh+t1g6+AzXe+oXu4+v2PZ+zRrE). | — |
| Consumer educational materials (native Spanish) | Spanish-language government brochures, NGO pamphlets, and web-based health coaching materials^105–116^. | — |
| Consumer educational materials (translated from English) | Evidence-based English health guides^117–124^ translated into Spanish using Google Translate v2. | — |
| TEDx transcripts | Spanish-language TEDx transcripts from the mTEDx corpus^45^. | salud, medicina, prevención, cuidado, ejercicio, cuerpo, dieta, alimentación, hábitos, hábitos saludables, fortalecer, recuperación, autocuidado, mente, emocional, salud mental, ansiedad, estrés, resiliencia, terapia, bienestar, motivación, respiración, sueño, dolor |

**Supplementary Table 3.** The dataset used to train MHC-Coach-ES in phase 2 consists of 3,268 expert-written prompt–completion pairs aligned with the Transtheoretical Model of behavior change and translated into Spanish (≈ 0.23 million BPE tokens). This dataset was used for instruction-style fine-tuning and was partitioned into 90% training and 10% validation splits.

| **Metric** | **Train** | **Validation** | **Total** |
| --- | --- | --- | --- |
| Examples (prompt–completion pairs) | 2,909 | 326 | 3,268 |
| Tokens (BPE) | 207,003 | 23,411 | 230,414 |

**Supplementary Table 4.**

Illustrative prompts and single example outputs for each message-generation strategy. Each row shows the input prompt and one representative message produced by a given generation strategy for the Maintenance stage of the TTM. The Base and Fine-tuned conditions use an identical 20-word stage-specific instruction. Few-shot prompting uses similar instructions with a system role, and three stage-matched example messages from the crowdsourced dataset ^59^. These expert message examples were excluded from subsequent quantitative message analyses. RAG prepends the instruction with the k = 2 most relevant text chunks automatically retrieved from the same expert corpus (only the first few lines of the first chunk are shown for brevity; total retrieved context ≈ 2,000 tokens). Expert messages are drawn from the crowdsourced dataset^59^, and were not produced by an LLM.

| **Generation Method** | **Prompt** | **Prompt Translated to English** | **Example Output Message** | **Output Translated to English** |
| --- | --- | --- | --- | --- |
| Base | Redacta cinco mensajes motivacionales en español, cada uno con exactamente 20 palabras, para una persona en la etapa de preparación del cambio de ejercicio.  Esta persona está lista para comenzar a hacer ejercicio en los próximos 30 días y ha empezado a dar pequeños pasos.  Usa un tono tranquilo, alentador y profesional, como el de un asesor de salud.  Escribe con lenguaje claro, natural y apropiado para una persona con nivel de lectura de sexto grado. Cada mensaje debe aplicar una técnica distinta. | Write five motivational messages in Spanish, each with exactly 20 words, for a person in the preparation stage of changing their exercise habits.  This person is ready to begin exercising in the next 30 days and has begun taking small steps.  Use a calm, encouraging, and professional tone, like that of a health coach.  Write in clear, natural language appropriate for a person with a sixth-grade reading level. Each message should use a different technique. | ¡Estás haciendo un gran trabajo! Cada pequeño paso hacia adelante es un logro. Sigue adelante, ¡tú puedes hacerlo! | You're doing a great job! Every small step forward is an achievement. Keep going, you can do it! |
| Fine-tuned (MHC-Coach) | Redacta cinco mensajes motivacionales en español, cada uno con exactamente 20 palabras, para una persona en la etapa de precontemplación del cambio de ejercicio.  Esta persona no planea comenzar a hacer ejercicio en los próximos seis meses y no considera su comportamiento actual como un problema.  Usa un tono tranquilo, alentador y profesional, como el de un asesor de salud.  Escribe con lenguaje claro, natural y apropiado para una persona con nivel de lectura de sexto grado. Cada mensaje debe aplicar una técnica distinta. | Write five motivational messages in Spanish, each with exactly 20 words, for a person in the precontemplation stage of exercise change.  This person does not plan to start exercising in the next six months and does not consider their current behavior a problem.  Use a calm, encouraging, and professional tone, like that of a health coach.  Write with clear, natural language appropriate for a person with a sixth-grade reading level. Each message should use a different technique. | Tu salud es muy importante para ti. Sin embargo, no haces ejercicio. ¿Cómo puedes cuidarte mejor? El ejercicio puede ayudarte a sentirte más sano. | Your health is very important to you. Yet, you don't exercise. How can you take better care of yourself? Exercise can help you feel healthier. |
| Few-shot | Redacta cinco mensajes motivacionales en español, cada uno con exactamente 20 palabras, para una persona en la etapa de acción del cambio de ejercicio.  Esta persona ha comenzado recientemente a hacer ejercicio (en los últimos seis meses) y está construyendo una nueva rutina saludable.  Usa un tono tranquilo, alentador y profesional, como el de un asesor de salud.  Escribe con lenguaje claro, natural y apropiado para una persona con nivel de lectura de sexto grado. Cada mensaje debe aplicar una técnica distinta.  Ejemplos:  Si continúas, ¡estos ejercicios te resultarán mucho más fáciles en el futuro!  Ya empezaste a hacer ejercicio, ¡sigue así! ¿Te das cuenta de cómo mejora tu bienestar mental?  Si sigues haciendo ejercicio con regularidad, puedes darte un capricho con algún dulce sin sentirte culpable. | You are a health coach trained in the Transtheoretical Model of Behavior Change. Write five motivational messages in Spanish, each with exactly 20 words, for a person in the action stage of exercise change.  This person has recently started exercising (within the last six months) and is building a new, healthy routine.  Use a calm, encouraging, and professional tone, like that of a health coach.  Write in clear, natural language appropriate for a sixth-grade reader. Each message should use a different technique.  Examples:  If you continue, these exercises will be much easier in the future!  You've already started exercising—keep it up! Do you notice how your mental well-being improves?  If you continue exercising regularly, you can treat yourself to a sweet treat without feeling guilty. | Ahora eres una persona que se cuida y se ama, y eso se refleja en tus elecciones saludables y en tu compromiso con el ejercicio. | You are now a person who cares for and loves yourself, and that is reflected in your healthy choices and your commitment to exercise. |
| RAG | ###CONTEXTUAL INFORMATION###  Redacta cinco mensajes motivacionales en español, cada uno con aproximadamente 10 palabras, para una persona en la etapa de Acción del cambio del cambio de ejercicio.\nCada mensaje debe aplicar una técnica distinta: Relaciones de Ayuda, Autoliberación, Gestión del Refuerzo, el Contracondicionamiento, Autoliberación.  […rest of retrieved chunks / ~3 000 tokens total …]  ################################  "Redacta cinco mensajes motivacionales en español, cada uno con exactamente 20 palabras, para una persona en la etapa de acción del cambio de ejercicio.  Esta persona ha comenzado recientemente a hacer ejercicio (en los últimos seis meses) y está construyendo una nueva rutina saludable.  Usa un tono tranquilo, alentador y profesional, como el de un asesor de salud.  Escribe con lenguaje claro, natural y apropiado para una persona con nivel de lectura de sexto grado. Cada mensaje debe aplicar una técnica distinta." | ###CONTEXTUAL INFORMATION###  Write five motivational messages in Spanish, each approximately 10 words long, for a person in the Action stage of the change exercise. Each message should apply a different technique: Helping Relationships, Self-Release, Reinforcement Management, Counterconditioning, Self-Release.  […rest of retrieved chunks / ~3 000 tokens total …]  ################################  five motivational messages in Spanish, each with exactly 20 words, for a person in the action stage of exercise change.  This person has recently started exercising (within the last six months) and is building a new, healthy routine.  Use a calm, encouraging, and professional tone, like that of a health coach.  Write with clear, natural language appropriate for a person with a sixth-grade reading level. Each message should use a different technique." | Recuerda que cada pequeño logro es un paso hacia adelante, no te desanimes, sigue adelante y verás los resultados. | Remember that every small achievement is a step forward. Don't get discouraged, keep going and you'll see the results. |
| Expert |  |  | Empezar a hacer ejercicio suele ser la parte más difícil, ¡y eso ya lo has conseguido! A partir de ahora todo será más fácil. | Starting to exercise is often the hardest part, and you've done it! From now on, everything will be easier. |

***S.1 MHC-Coach-ES Fine-Tuning Dataset Preprocessing***

**S.1.1 MHC-Coach-ES Corpus Cleaning and Chunking**

All raw text was preprocessed to ensure standardized chunking and normalization of characters. First, the text was normalized to Unicode NFKC and repaired with the ftfy Python library (version 6.3.1) to correct mojibake and malformed accent marks. Control characters and excess whitespace were then removed, and curly quotation marks were replaced with straight equivalents to stabilize downstream tokenization. The cleaned text was segmented on Spanish sentence-ending punctuation ( . ? ! ¡ ¿ ) and concatenated into chunks no longer than 500 tokens (measured by the LLaMA-3-70B SentencePiece tokenizer).

**S.1.2 MHC-Coach-ES Data Split**

After preprocessing, the SHMC comprised 2.21 M tokens. For each source category, document chunks were shuffled using a fixed random seed and divided into 90% training and 10% validation splits, using stratified sampling to preserve the category-wise distribution. The held-out validation set was used exclusively for hyperparameter selection. Detailed token counts for each source category and split are provided in **Supplementary Table 2** and **Supplementary Table 3**.

***S.2 Generation Methods***

To contextualize the linguistic and stylistic properties of MHC-Coach-ES, we compared it against other feasible approaches for generating proactive, single-turn coaching messages in Spanish. We generated twenty messages, four per TTM stage, using five methods: (i) the base Llama-3-70B-Instruct model, (ii) MHC-Coach-ES, (iii) few-shot prompting, (iv) retrieval-augmented generation (RAG)^52^, and (v) translated human-expert written message.

Across all generation methods, we used a shared base task prompt that specified the target Transtheoretical Model (TTM) stage, included a concise description of the TTM stage, and requested a concise (approximately 20-word) motivational message in Spanish. Method-specific context, such as example messages in the few-shot condition or retrieved reference text in the RAG condition, was added on top of this common instruction to reflect standard implementations of each approach.

**S.2.1 Llama-3-70B-Instruct base model**

The unmodified Llama-3-70B-Instruct model was prompted with the shared base task instruction, which included a brief description of the target TTM stage and requested a concise (approximately 20-word) motivational message in Spanish tailored to that stage.

**S.2.2 Few-shot prompting**

We leveraged the same stage-specific task description used in the baseline, along with three stage-matched demonstration messages. Examples were drawn from the translated human-expert corpus (expert message in T-TTM) but held out from the subset used for quantitative comparisons. The concatenated prompt was provided to the unmodified Llama-3-70B-instruct model.

**S.2.3 Retrieval-augmented generation (RAG)**

We constructed a Spanish reference corpus comprising the Translated TTM Coaching Dataset (T-TTM), containing 3,268 translated stage-labeled expert messages, together with the exercise-science excerpts from the Spanish Health & Motivation Corpus (SHMC) described in Methods Section M.1.1 and M.1.2. This combined corpus was embedded using the BAAI/bge-small-en-v1.5 encoder^103^ and indexed with LlamaIndex v0.10.38^104^. Documents were split into contiguous 1,000-token chunks without overlap, and an embedding was computed for each chunk. At inference, a message that requested for a motivational message for a specific stage of change was translated to Spanish then embedded usingBAAI/bge-small-en-v1.5 encoder, and three chunks from the reference corpus with the highest cosine similarity to the query were retrieved. These three segments were prepended to the same instruction prompt used in the baseline condition (see S.2.1), and the full context was provided to the unmodified Llama-3-70B-instruct model for generation.

**S.2.4 Expert Messages**

Twenty expert-written messages were randomly sampled from the crowd-sourced corpus used in the training data and described in Section M.4.3.

Example prompts and generated messages for each generation approach are provided in **Supplementary Table 4**.

***S.3 Linguistic Analysis***

**S.3.1 Corpus and preprocessing**

We performed a systematic comparison of the linguistic and stylistic properties of the messages produced in each generation technique described in Methods Section M.3.1. Each of the five generation methods produced 20 messages, yielding a corpus of 100 messages.

All analyses were run in Python 3.12. Linguistic annotation (tokenization, lemmatization, part-of-speech tagging, dependency parsing, and named entities) used spaCy 3.7.4 with the es_core_news_sm model. Sentiment was computed with pysentimiento 0.7.3 (Spanish), and readability with Textstat 0.7.3 using Spanish indices when available.

**S.3.2 Feature definition and extraction**

The selected features reflect complementary dimensions of message quality, including length, lexical richness, emotional tone, actionability, temporal specificity, and readability. Each feature is defined below, along with the method used for its extraction.

*S.3.2.1 Word Count*

Word count was calculated as the total number of alphabetic tokens in each message, with numerals and punctuation excluded. Tokenization and filtering were performed using spaCy.

*S.3.2.2 Continuous Sentiment Score*

A transformer-based Spanish sentiment classifier was used via pysentimiento (version 0.7.3), which returns posterior probabilities over the labels NEG, NEU, POS. For each message we define a continuous sentiment index, S=P(POS)−P(NEG)∈[−1,1], so that larger values indicate a more positive tone and more negative values a more negative tone.

*S.3.2.3 Temporal Anchoring*

This feature captures whether a message is explicitly anchored in time. Messages were assigned a value of 1 if any of the following criteria were met:

(i) a Spanish named-entity recognizer identified a temporal entity (DATE or TIME), implemented using spaCy;
 (ii) the text matched an entry in a curated Spanish lexicon of temporal expressions following case folding and diacritic removal (Unicode NFKD normalization); or
 (iii) the text matched regular-expression patterns encoding common Spanish temporal constructions, including frequencies (e.g., “al/a la/por día, semana, mes, año”), counts (e.g., “N veces por …”), durations (e.g., “durante X días/meses/años,” “desde hace X semanas,” “hace X …”), weekdays (lunes–domingo), and clock times (e.g., “a las HH:MM”).

Messages that did not satisfy any criterion were assigned a value of 0. This composite approach increased recall relative to named-entity recognition alone, capturing expressions such as “150 minutos a la semana” and “durante más de seis meses.” Regular-expression matching was implemented using Python’s standard regex engine, with text normalization performed via the unicodedata module.

*S.3.2.4 Type–Token Ratio*

Lexical diversity was computed as the ratio of unique alphabetic tokens to total alphabetic tokens after normalizing tokens to lowercase and removing diacritics.

*S.3.2.5 Exclamation Point Count*

We quantified the total number of exclamatory characters in the message, counting both the inverted Spanish mark “¡” and the standard mark “!”. Paired marks (e.g., “¡…!”) contributed two to the total count, and contiguous repeats (e.g., “!!!”) were counted per character.

*S.3.2.6 Readability*

Readability was estimated using Spanish formulas implemented in Textstat (see M.3.2). We employed the Fernández–Huerta index, where higher scores indicate greater ease of reading.
